# Is sickle cell disease a risk factor for severe COVID-19 : a multicenter national retrospective cohort

**DOI:** 10.1101/2020.12.30.20249053

**Authors:** Abdulkarim Abdulrahman, Mohammed Wael, Fajer AlAmmadi, Zahra AlMosawi, Reem Alsherooqi, Manal AlSayed, Nitya Kumar, Manaf AlQahtani

**Author notes:** Corresponding author: Manaf AlQahtani.

## Abstract

**Introduction:** Coronavirus disease (COVID-19) caused by the novel coronavirus SARS-CoV-2 is an infectious disease which has evolved into a worldwide pandemic. Growing evidence suggests that individuals with pre-existing comorbidities are at higher risk of a more serious COVID-19 illness. Sickle cell disease (SCD) is an inherited hemoglobinopathy which increases the susceptibility to infections and as a consequent has higher risks of morbidity and mortality.

The impact of COVID-19 on SCD patients could lead to further increase in disease severity and mortality. Studies that examine the effect of SCD on COVID-19 outcomes are lacking. This study aims to determine whether SCD is a risk factor for severe COVID-19 infection in regards to the requirement of non-invasive ventilation/high flow nasal cannula (NIV/HFNC), mechanical intubation (MV) or death.

**Methods:** Retrospective cohort study which included COVID-19 patients admitted to four Ministry of Health COVID-19 treatment facilities in Bahrain during the period of 24, February 2020, to 31, July 2020. All SCD patients with COVID-19 were included and compared to randomly selected non-SCD patients with COVID-19. Data for the selected patients were collected from the medical records. Multivariate logistic regression models were used to control for confounders and estimate the effect of SCD on the outcomes.

**Results:** A total of 1,792 patients with COVID-19 were included; 38 of whom were diagnosed with SCD as well. In the SCD group, one (2.6%) patient required NIV/HFNC, one (2.6%) required MV and one (2.6%) death occurred. In comparison, 56 (3.2%) of the non-SCD patients required NIV/HFNC, 47 (2.7%) required MV and death occurred in 58 (3.3%) patients. Upon adjusting for confounders, SCD had an odds ratio of 1.847 (95% CI: 0.39 – 8.83; p=0.442).

**Conclusion:** Our results indicate that SCD is not a risk factor for worse disease outcomes in COVID-19 patients.

## Introduction

The first case of the novel Coronavirus (SARS-CoV-2) was diagnosed in Wuhan, China in December 2019. Since then, the virus has spread drastically causing a pandemic that resulted in more than 81 million cases to date and more than 1.7 million deaths worldwide^1^. This led to global health and economic crises in multiple countries, many of which are projected to face long-lasting repercussions^2^.

The illness caused by the SARS-CoV-2 virus varies in its symptomatology and can range from being asymptomatic^3^ to severe illness leading to acute respiratory distress syndrome and eventually, death^4^. Several studies have shown an association between higher rates of hospitalization and severe illness with advanced age, male gender, heart failure, diabetes mellitus, severe asthma, high body mass index, chronic kidney disease, chronic pulmonary disease and malignancy^5–8^. Additional factors that are suggestive of more serious COVID-19 complications include admission oxygen saturation of <88%, troponin-I level >1 ng/mL, C-reactive protein level >200 mg/L, and D-dimer levels >2500 ng/mL^6^. COVID-19 has been shown to increase the risk of thromboembolism which can lead to significant morbidity and mortality^9^.

Sickle cell disease (SCD) is an inherited group of disorders characterized by the presence of Hemoglobin S (HbS), either from homozygosity for the sickle mutation in the beta globin chain of hemoglobin (HbSS) or from compound heterozygosity of a sickle beta globin mutation with another beta globin mutation (e.g., Sickle-Beta Thalassemia). Hemoglobinopathies, sickle cell disease in particular, are commonly found in the Kingdom of Bahrain^10^. In 1995, a large study involving the hospitalized population of Bahrain which included 5,503 neonates and 50,695 non-neonates, reported the prevalence of SCD as 2.1% and HbS trait as 18.1% in neonates, and 10.4 % SCD in non-neonatal patients^11^.

A hereditary blood disorder prevention awareness campaign was started in 1985 which drastically reduced the incidence of hereditary blood disorders in Bahrain^12^. In May 2007, newborn screening for SCD and thalassemia was established^13^. The program was offered to all newborns, where cord blood was tested for hemoglobinopathies. A total of 38,940 newborns were screened between 2007 and 2010. The incidence of SCD in newborns was found to be 0.7%, 0.6% and 0.4% in the years 2008, 2009 and 2010 respectively in comparison to 2.1% in 1985^12^.

Patients with SCD are at a higher risk of morbidity & mortality. As per the nature of the disease, they are predisposed to developing thrombotic events. These include vaso-occlusive crises, cerebrovascular accidents, acute sickle hepatic crises, acute hepatic sequestration and renal disease. Hemolytic crises, aplastic crises, infective episodes and priapism are also amongst the complications seen in these patients. This can explain why such patients tend to have a reduced life expectancy when compared to the general population^14,15^. Severe complications such as acute chest syndrome^16^ and multiple organ failure^17^ might be catastrophic and eventually lead to death.

Mortality rates for SCD patients admitted to intensive care units in Bahrain were 12.7%. This was slightly less than the overall mortality in other centers worldwide which was 17.2%.^18^

Critically ill patients can develop cytokine storm, progressive endothelial activation and disseminated intravascular coagulation (DIC) resulting in multi-organ failure^19^.

Patients with SCD often have multiple comorbidities including chronic lung disease. Viral infection can trigger acute vaso-occlusive crises in SCD patients, especially acute chest syndrome which is associated with high mortality rates^20^. In this setting of multi-organ dysfunction, in particular chronic lung damage, COVID-19 could easily trigger ACS and multi-organ failure. Underlying endothelial dysfunction and abnormal expression of procoagulants such as tissue factor could also place SCD patients at a greater risk^21^.

Hence, the combination of COVID-19 and SCD is hypothesized to cause an increase in disease severity and mortality. Studies that examine the effect of SCD on COVID-19 outcomes are lacking. Given the morbidity and mortality associated with sickle cell disease patients, it is predicted that these patients might have unfavorable outcomes when infected with the SARS-Cov-2 virus.

This study aims to determine if SCD is a risk factor for severe COVID-19 through comparing severity and outcomes of COVID-19 cases.

### Objectives

1. Compare the odds of non-invasive ventilation, intubation or death in SCD cases and controls
2. Compare the odds of requiring oxygenation on presentation or admission in SCD cases versus controls

## METHODS

### Study design and setting

The study cohort included COVID-19 patients admitted to Bahrain’s Ministry of Health COVID-19 treatment facilities during 24 February 2020 till 31 July 2020. The four participating institutions included: Ebrahim bin Khalil Kanoo COVID-19 Centre, Salmaniya Medical Complex COVID-19 Centre, Hereditary Blood Disorder Centre (HBDC) COVID-19 Centre and Jidhafs COVID-19 Centre. All SCD patients diagnosed with COVID-19 were included. Patients without SCD were sampled randomly from this COVID-19 study cohort.

### Outcomes

#### Primary outcome

The primary endpoint was the requirement of non-invasive ventilation, intubation or death.

#### Secondary outcome

The requirement for oxygenation on presentation or admission

### Diagnosis

The following methods were used to diagnose COVID-19 and sickle cell disease in our study

#### COVID-19

All patients were confirmed to be infected by SARS-CoV-2 using a PCR test of a nasopharyngeal sample. The PCR test was conducted using Thermo Fisher Scientific (Waltham, MA) TaqPath 1-Step RT-qPCR Master Mix, CG on the Applied Biosystems (Foster City, CA) 7500 Fast Dx RealTime PCR Instrument. The assay used and targeted the E gene. If the E gene was detected, the sample was then confirmed by RdRP and N genes. The E gene Ct value was reported and used in this study. Ct values >40 were considered negative. Positive and negative controls were included for quality control purposes.

#### Sickle Cell Disease

SCD status of patients was obtained from the medical records. All patients marked as SCD within the study time frame were included. Patients with sickle cell trait were not considered as SCD. The diagnosis of SCD is made through hemoglobin electrophoresis and confirmed by a positive sickling test result.

### Patient Selection

A full list of COVID-19 patients in these study periods and their SCD status were extracted from the electronic medical records. The list was imported into the statistical package. All COVID-19 patients with SCD as indicated in the EMR, were included. A total of 1850 non-SCD patients with COVID-19 were randomly sampled. Ninety-six patients were excluded due to duplicates, non hospitalized or due to lack of data. Data was collected for a total of 1754 patients.

### Data sources and variables assessed

We obtained data from the national electronic medical records database in Bahrain, “I-SEHA”. Since this EMR database stores information as unstructured data, the study participants’ information was manually extracted by five physicians, assisted by ten senior medical students. This team reviewed all the cases and collated the information into a structured dataset for this study. The data gathered included patients’ demographic details, vital signs, laboratory test results, medication lists, past medical history, SCD status, clinical severity scale (as seen in the supplementary table attached in the appendix), oxygenation requirement on admission, the ratio of the oxygen saturation to the fraction of inspired oxygen (SpO2:FiO2) at admission, requirement of ventilator use and outcomes.

### Statistical analysis

The distribution of treatment groups was summarized. Bivariate associations between the sickle cell status groups and the measured patient characteristics were analyzed using Chi-squared (χ2) tests for categorical variables and t-test for continuous variables. We assessed endpoints and their associations with both groups.

Multivariable logistic regression models were used to estimate the relationships between SCD and the composite primary and secondary endpoints. These models included demographic factors, clinical factors and medications.

The STATA software, version 15.1, was used to execute the statistical analyses, (StataCorp. 2017. Stata Statistical Software: Release 15. College Station, TX: StataCorp LLC.).

### Ethical approval

The protocol and manuscript for this study were reviewed and approved by the National COVID-19 Research Committee in Bahrain. This committee has been jointly established by the Bahrain Ministry of Health and Bahrain Defence Force Hospital research committees in response to the pandemic, to facilitate and monitor COVID-19 research in Bahrain. All methods and retrospective analysis of data was approved by the National COVID-19 Research and Ethics Committee, and carried out in accordance with the local guideline and ethical guidelines of the Declaration of Helsinki 1975. All data used in this study was collected as part of normal medical procedures.

Informed consent was waived by the National COVID-19 Research and Ethics Committee for this study due to its retrospective and observational nature and the absence of any patient identifying information.

## Results

1792 cases were included. During the study period from 24^th^ of February 2020 until 31^st^ of July 2020; a total of 38 patients diagnosed with Sickle Cell Disease (SCD) were admitted with SARS Cov-2 infection. The outcomes of these patients were compared to a randomly selected control group of 1754 patients diagnosed with SARS Cov-2 during the same study period. There were a number of significant differences between the two groups (Table 1, 2, 3). The mean age of the SCD group was 35.5 years in comparison to 46.2 in the non-SCD group. 28.9% of the SCD group were males while 59.6% were males in the non-SCD group. All the SCD cases were Bahraini nationals, compared to 55.7% in the non-SCD group. Mean age, proportion of males, Bahraini nationals and G6PD deficiency status were significantly different in SCD cases and non-cases. It was also evident that diabetes and hypertension were more common in the control group. In the SCD group, 13.2% were diabetic compared to 29.2% in the non-SCD group, similarly, 13.2% of the SCD cases were hypertensive compared compared to 30.1% of the non-SCD group. On the other hand, G6PD deficiency was more prevalent within the SCD group (44.7%) while only 9% of the non-SCD patients had this deficiency. All these differences were statistically significant with p-value <0.05. The prevalence of cardiovascular disease was similar across both groups (10% and 10.5%).

**Table 1:** Baseline characteristics of non-SCD and SCD groups

| Factor | Level | Non-SCD | SCD | p-value |
| --- | --- | --- | --- | --- |
| N |  | 1754 | 38 |  |
| Age – mean ± SD |  | 46.2 ± 16.9 | 35.5 ± 18.7 | <0.001 |
| Male |  | 1046 (59.6%) | 11 (28.9%) | <0.001 |
| Bahraini |  | 977 (55.7%) | 38 (100.0%) | <0.001 |
| Presence of comorbidities (Other than SCD) | Yes | 863 (49.2%) | 22 (57.9%) | 0.29 |
| Number of comorbidities (Other than SCD) | 0 | 891 (50.8%) | 16 (42.1%) | 0.005 |
|  | 1 | 422 (24.1%) | 17 (44.7%) |  |
|  | 2 | 258 (14.7%) | 0 (0.0%) |  |
|  | 3+ | 183 (10.4%) | 5 (13.2%) |  |
| G6PD Deficiency |  | 158 (9.0%) | 17 (44.7%) | <0.001 |
| Diabetes Mellitus |  | 512 (29.2%) | 5 (13.2%) | 0.031 |
| Cardiovascular Disease (CVD) |  | 176 (10.0%) | 4 (10.5%) | 0.92 |
| Hypertension |  | 528 (30.1%) | 5 (13.2%) | 0.024 |
| Asthma |  | 78 (4.4%) | 2 (5.3%) | 0.81 |
| Chronic Obstructive Pulmonary Disease (COPD) |  | 7 (0.4%) | 0 (0.0%) | 0.70 |
| Chronic Kidney Disease (CKD) |  | 77 (4.4%) | 2 (5.3%) | 0.80 |
| Other Chronic Lung Disease (not asthma nor COPD) |  | 9 (0.5%) | 0 (0.0%) | 0.66 |

**Table 2:** Clinical characteristics of non-SCD and SCD groups

| Factor | Level | Non-SCD | SCD | p-value |
| --- | --- | --- | --- | --- |
| Fever (>38C) |  | 385 (21.9%) | 16 (42.1%) | 0.003 |
| Cough |  | 735 (41.9%) | 18 (47.4%) | 0.50 |
| Chest Pain |  | 153 (8.7%) | 6 (15.8%) | 0.13 |
| Shortness of Breath |  | 354 (20.2%) | 6 (15.8%) | 0.50 |
| Loss of smell |  | 53 (3.0%) | 3 (7.9%) | 0.088 |
| Loss of taste |  | 50 (2.9%) | 4 (10.5%) | 0.006 |
| Diarrhea |  | 101 (5.8%) | 3 (7.9%) | 0.58 |
| Nausea or Vomiting |  | 91 (5.2%) | 2 (5.3%) | 0.98 |
| Body pain |  | 266 (15.2%) | 12 (31.6%) | 0.006 |
| Heart Rate on admission: bpm - mean ±SD | | 86.6 ±14.2 | 91.8 ±15.1 | 0.027 |
| SBP on admission: mmHg - mean ± SD | | 130.6 ±18.7 | 121.4 ±15.0 | 0.004 |
| DBP on admission: mmHg - mean ± SD | | 77.3 ± 11.7 | 70.9 ±11.1 | 0.001 |
| Respiratory rate on admission: breath per minute - mean ± SD | | 20.1 ± 6.9 | 20.2 ± 1.5 | 0.97 |
| Elevated ALT>40U/L |  | 398 (24.6%) | 10 (27.8%) | 0.66 |
| Elevated Creatinine |  | 166 (9.5%) | 3 (7.9%) | 0.74 |
| CXR findings | Pneumonia | 515 (34.4%) | 16 (44.4%) | 0.21 |
|  | Normal | 981 (65.6%) | 20 (55.6%) |  |
| Hypotension (SBP<90mmHg or DBP<60mmHg) on admission |  | 72 (4.1%) | 5 (13.2%) | 0.006 |
| Tachypnea (RR>22) on admission |  | 49 (2.8%) | 0 (0.0%) | 0.30 |
| Disease severity on admission | On room air | 1542 (87.9%) | 33 (89.2%) | 0.97 |
|  | On oxygen support (nasal cannula/ face mask) | 201 (11.5%) | 4 (10.8%) |  |
|  | On NIV/HFNC | 8 (0.5%) | 0 (0.0%) |  |
|  | Mechanical ventilation | 3 (0.2%) | 0 (0.0%) |  |
| Oxygenation on admission |  | 212 (12.1%) | 4 (10.5%) | 0.77 |

**Table 3:** COVID-19 related Medications received during hospital stay in non-SCD and SCD groups

| Medication | Non-SCD | SCD | p-value |
| --- | --- | --- | --- |
| Hydroxychloroquine | 531 (30.3%) | 6 (15.8%) | 0.054 |
| Azithromycin | 430 (24.5%) | 15 (39.5%) | 0.035 |
| Kaletra | 318 (18.1%) | 6 (15.8%) | 0.71 |
| Ribavirin | 267 (15.2%) | 6 (15.8%) | 0.92 |
| Steroids | 198 (11.3%) | 6 (15.8%) | 0.39 |
| Tocilizumab | 81 (4.6%) | 1 (2.6%) | 0.56 |
| Convalescent Plasma Transfusion | 65 (3.7%) | 3 (7.9%) | 0.18 |

Results also demonstrated that patients who had SCD had additional comorbidities. In the SCD group, 44.7% were diagnosed with at least one other comorbidity and 13.2% had three or more other co-morbidities in contrast to the 24.1% and 10.4% respectively in the non-SCD group.

The characteristics of the studied sample are summarized in Tables 1 to 3

Among the 33 (71.1%) of the symptomatic SCD patients, cough was the most common symptom (47.4%), followed by fever (42.1%). On the other hand, 1108 (63.2%) of non-SCD patients were symptomatic with similar common symptoms of cough (41.9%) and fever (21.9%).

Of the SCD group, 33 (89.2%) patients were admitted on room air, and 4 (10.8%) patients were admitted with oxygen support. None of the SCD patients admitted required further support on admission. Among the non-SCD patients, 1542 (87.9%) were admitted on room air, 201 (11.5%) on oxygen support, 8 (0.5%) on NIV/HFNC and 3 (0.2%) required mechanical ventilation (MV).

### Disease outcomes

In the SCD group, one (2.6%) patient required NIV/HFNC, one (2.6%) required MV and one (2.6%) death occurred. In comparison, 56 (3.2%) of the non-SCD patients required NIV/HFNC, 47 (2.7%) required MV and mortality was seen in 58 (3.3%). The outcomes are summarized in Table 4.

**Table 4:** Outcomes in non-SCD and SCD groups

| Factor | Non-SCD | SCD | p-value |
| --- | --- | --- | --- |
| NIV/HFNC/MV or Death | 109 (6.2%) | 3 (7.9%) | 0.67 |
| NIV/HFNC | 56 (3.2%) | 1 (2.6%) | 0.85 |
| Intubation | 47 (2.7%) | 1 (2.6%) | 0.99 |
| Death | 58 (3.3%) | 1 (2.6%) | 0.82 |

### Primary outcome

The primary outcome of the study is to evaluate the composite outcome of the requirement for ventilation (invasive or noninvasive) or death or mortality (Table 5). The total number of patients who developed the primary outcome in both groups is 112/1792 (6.3%), of which 3/38 (7.9%) were in the SCD group and 109/1754 (6.2%) in the non-SCD group. The unadjusted odds ratio for SCD patients developing the primary outcome was 1.29 (95% CI: 0.39 – 4.27; p=0.67). Upon adjusting for confounders, we found the odds ratio to be 2.06 (95% CI: 0.43 – 9.95; p= 0.37). Table 5 summarizes these.

**Table 5:** Risks for developing the primary outcome

| Analysis | Ventilation or Death | p-value |
| --- | --- | --- |
| No. of events/no. of patients at risk (%) | 112/1792 (6.3%) | - |
| SCD | 3/38 (7.9%) | - |
| Non-SCD | 109/1754 (6.2%) | - |
| Crude analysis – odds ratio (95%CI) | 1.29 (0.39 – 4.27) | 0.67 |
| Multivariable analysis* – odds ratio (95%CI) | 1.847 (0.39 – 8.83) | 0.442 |
| *covariates included were age, gender, hypertension, chronic kidney disease, severity scale on admission hydroxychloroquine, azithromycin, steroids, tocilizumab, plasma transfusion |  |  |

### Secondary outcome

The secondary outcome of the study was the requirement of oxygenation on presentation (Table 6). The total number of patients who required oxygenation upon presentation was 12.1% (216/1792), of which 7.9% (4/38) were from the SCD group and 12.1% (212/1754) were from the non-SCD group. The unadjusted odds ratio was found to be 0.85 (95% CI: 0.30 – 2.44, p=0.77). The adjusted odds ratio was found to be 1.39 (95% CI: 0.41 – 4.6; p= 0.59).

**Table 6:** Risks for developing the secondary outcome

| Analysis | Oxygenation | p-value |
| --- | --- | --- |
| No. of events/no. of patients at risk (%) | 216/1792 (12.1%) | - |
| SCD | 4/38 (7.9%) | - |
| Non-SCD | 212/1754 (12.1%) | - |
| Crude analysis – odds ratio (95%CI) | 0.85 (0.30 – 2.44) | 0.77 |
| Multivariable analysis* – odds ratio (95%CI) | 1.39 (0.41 – 4.61) | 0.59 |
| *covariates included were age, male, Bahraini nationality, hypertension, chronic kidney disease, COPD, symptoms, fever, cough shortness of breath, heart rate, diastolic blood pressure |  |  |

Complete output and additional details on the models are available in the Appendix.

## Discussion

In this cohort study, SCD was not observed to be significantly associated with developing the primary composite outcome (requirement of NIV/HFNC/MV or death) in COVID-19 patients. SCD patients also did not have significantly higher odds of requiring oxygenation upon admission to the hospital as compared to non-SCD patients.

Upon analyzing the baseline characteristics of both groups, it was evident that there were significant differences in multiple domains. The non-SCD population was on average older than the SCD group. The younger mean age in SCD patients can be explained by the nature of the disease resulting in a reduced life expectancy^22^. Age has been reported to be associated with severe disease progression in COVID-19^23^. Most studies referred to age more than 60 years to increase the risk of worse outcomes. A study aiming to quantify the isolated effect of age on the severity of COVID-19 outcomes concluded a minimal influence after adjusting for important age-dependent risk factors (for example: diabetes, hypertension, cardiovascular disease etc.)^24^. On that account, although our study showed a difference of 10 years between the two groups, this might not have an actual effect on disease outcomes (since mean age was 46 and 35 years in the non-SCD and SCD groups, respectively). Regardless, age was adjusted for in the analysis conducted in this study.

All SCD patients were Bahraini nationals, compared to 55% of the patients in the non-SCD group. SCD is a genetic disorder that affects around 75,000 Bahrainis^11,25^. Bahrain has a wide range of nationalities and around 50% of Bahrain’s population comprises expatriate workers (with the majority being from Asian nationalities)^25^. SCD is a very rare disease within these countries^26,27^ and hence this can explain the nationality difference observed. In addition, the majority of expatriate workers are male majority^25^ and this accounted for the male predominance observed in the non-SCD group. Moreover this difference in gender distribution can be explained by the increased female prevalence of COVID-19 patients with hemoglobinopathies. A multicenter study by *de Sanctis et. al*^28^ and another study by *Panepinto et al*^29^ established increased female prevalence (76.9% and 57% respectively) of COVID-19 infection among patients with hemoglobinopathies.

Among the comorbidities evaluated in all patients with COVID-19, results demonstrated that almost half of those with SCD were found to have G6PD deficiency in contrast to a minority in the control group. Some studies claim that G6PD deficiency incidence is more abundant in patients with SCD in comparison to the general population^30^. In addition, many studies have established the association of such inherited diseases due to the high rates of consanguineous marriages in the gulf region^31^. Further research on G6PD and COVID-19 outcomes is required to understand more on the topic.

On a different note, patients in the non-SCD group were found to be mostly comorbid with diabetes mellitus (DM) and hypertension (HTN), this could be attributed to the older age of the population^32^. The higher prevalence of DM and HTN increased the baseline risk of the non-SCD group to develop severe COVID-19 outcomes^5–7^.

One of the remarkable findings in this study was the significant variation in symptom presentation. Although the majority of patients in both groups did present with cough, SCD patients presented more expressively with fever. This may be attributed to the natural history of the disease where it creates an environment supporting infections^33^, although the presence of other causes of fever were not explored in this study.

COVID-19 infection has a broad clinical presentation; ranging from asymptomatic to severe forms of respiratory disease which can lead to death^34^. Few studies have been published studying the correlation between SCD and the severity of COVID-19 infection. A study by *Panepinto et al*^29^ explored COVID-19 disease outcome in SCD patients living in the United States during the period March – May 2020. It concluded that this group of patients had increased risk of severe disease outcome as well as an increased mortality with 69% hospitalization rate, 11% ICU admission rate and 7% mortality rate. These were higher when compared to COVID-19 outcomes in the general population from a similar time period where hospitalization rate was 14%, ICU admission rate 2.3% and mortality rate 5.4%^35^.

Moreover, within the United States African American community, COVID-19 patients with SCD had higher rates of death from COVID-19 than those without SCD, as reported by Lana Mucalo et al at the American Society of Hematology (ASH) virtual meeting. Their study indicated that this population had 6.2 times the risk for COVID-19-related mortality^36^.

Another study, however, by Ashima Singh et al, indicated that African American individuals with SCD had similar risk of COVID-19-related mortality as those without SCD, after balancing for age, gender and other pre-existing conditions. Their study did report that SCD imposes additional risk of severe COVID-19 illness and hospitalization^37^.

In our study, SCD patients diagnosed with COVID-19 infection did not have a significant increase in risk for worse disease outcomes in comparison to the non-SCD patients. Similarly, SCD patients with COVID-19 did not have increased risk for requirement of oxygenation upon admission as compared those without SCD.

We have previously reported the disease outcomes in a case series of all SCD patients infected with COVID-19 between February and April 2020 in Bahrain. Six patients with SCD had COVID-19: three remained asymptomatic; two had mild symptoms and only one patient required oxygen support. The SCD patients had a similar average length of stay when compared with non-SCD COVID-19 patients^38^. Other studies also showed similar results. *Chakravotry et al*^*3*9^ explored COVID-19 disease outcomes of 10 SCD patients in a tertiary hospital in the United Kingdom. The study demonstrated that most patients had mild disease with 80% not requiring oxygen and the remaining 20% requiring only a nasal cannula. Only one mortality case was described in a 54-year old female that had severe pre-existing lung disease and history of previous admissions to the intensive therapy unit within the past year. Likewise, *Faiz A et al*^40^ presented a case series of four SCD patients diagnosed with COVID-19 infection. Three cases had mild disease and only one patient required oxygenation. All cases were discharged without mortalities.

Another case series *by Ramachandran et al* established low morbidity and mortality with COVID-19 in SCD patients. Among the nine SCD patients, only one required ICU admission but without the need for intubation. All cases were discharged and no deaths were observed^41^.

It is important to consider the sample used for this study as it represents hospitalized cases only. It is possible that other SCD patients remained in home isolation and did not require hospitalization and hence not included. Nevertheless, this would have not affected the results of this study. If these cases were included, the results would have moved more towards the null hypothesis.

## STRENGTHS

The study has several strengths; it involved the majority of hospitals that provided acute care for hospitalized COVID-19 cases in Bahrain. Furthermore, our study included all SCD cases admitted in the study time period allocated at the involved centres. The data collection process was done manually, hence, all patients’ files were reviewed carefully and all documented details were collected. The study represents the only study conducted in the region to explore the relationship between SCD and COVID-19.

## LIMITATIONS

Since the study includes severe COVID-19 disease patients requiring hospitalization, inference on milder COVID-19 infections cannot be drawn from these results. Secondly, given the retrospective design, information that was not documented in EMR and was not available for analysis, could be potential confounders. These included: time from symptom onset, inflammatory markers and frequency of admission of SCD patients. This research did not study other relevant outcomes related to sickle cell disease including hemolysis, vaso-occlusive crisis, and other SCD crises. The genotype of sickle cell disease patient was not stratified as data on that wasn’t readily available.

## CONCLUSION

Our results showed no significant increased risk of adverse COVID-19 outcome in SCD patients as compared to non-SCD patients. Further studies on this topic are warranted in order to have conclusive evidence on SCD and the risk of COVID-19 outcomes.

## Data Availability

All the data for this study will be made available upon reasonable request to the corresponding author.

## Acknowledgements

We would like to express our gratitude towards our colleagues: Abdulla AlAwadhi, Marwa AlMadhi, Islam AlSayed, Jumana AlArayed, Sara Jaafar Mohammed, Aesha Khalid Sharif, Khadija Alansari, Ammar Kheyami, Mujtaba Mal Alla, Abdulla AlMuharraqi, Zeyad Mahmood, Narjis Ali AlSheala, Ola Husain AlHalwachi, Maryam Ghazi Alarayedh, and Amna Mohamed Buheiji who played an essential role in the data collection process related to this paper. Our thanks and appreciation goes to them for their dedication and hard work and we wish them all the best in the future. We thank all the participants of the study and hope they will lead a long healthy life. We extend our condolences to the families of the lives lost due to COVID-19.

## Disclosures

### Ethics approval and consent to participate

The study was approved by the National COVID-19 Research and Ethics Committee.

### Consent for publication

All authors gave their consent for publication.

### Author contributions

AA, MQ gathered the data and supervised the data collection team. AA and NK analyzed and interpreted the data. AA, FA, MW, RS, ZM, wrote the manuscript. MQ, MA, NK edited the manuscript. All authors reviewed and approved the final version of the manuscript. MQ is the guarantor of this work.

