## Appendix for "Is sickle cell disease a risk factor for severe COVID-19 : a multicenter national retrospective cohort"

### Primary outcome analysis: Requirement for NIV/HFNC/MV or Death

**Logistic regression: Crude**

| outcome | Coef. | | | St.Err. | t-value | | p-value | [95% Conf | | Interval] | Sig |
| --- | --- | --- | --- | --- | --- | --- | --- | --- | --- | --- | --- |
| Sickle cell disease | | 1.294 | | 0.789 | 0.42 | | 0.673 | 0.392 | | 4.273 |  |
| Constant | 0.066 | | | 0.007 | -27.44 | | 0.000 | 0.055 | | 0.080 | *** |
| Mean dependent var | | | 0.062 | | | SD dependent var | | | 0.242 | |  |
| Pseudo r-squared | | | 0.000 | | | Number of obs | | | 1792.000 | |  |
| Chi-square | | | 0.167 | | | Prob > chi2 | | | 0.683 | |  |
| Akaike crit. (AIC) | | | 841.743 | | | Bayesian crit. (BIC) | | | 852.725 | |  |
| **** p<0.01, ** p<0.05, * p<0.1* | | | | | | | | | | |  |

**Logistic Regression: Multivariate**

| Primary outcome | Coef. | | St.Err. | t-value | | p-value | [95% Conf | | Interval] | Sig |
| --- | --- | --- | --- | --- | --- | --- | --- | --- | --- | --- |
| Sickle Cell disease | 1.847 | | 1.474 | 0.77 | | 0.442 | 0.386 | | 8.826 |  |
| Age | 1.005 | | 0.012 | 0.42 | | 0.674 | 0.982 | | 1.029 |  |
| Male | 1.049 | | 0.324 | 0.16 | | 0.876 | 0.573 | | 1.923 |  |
| Hypertension | 2.778 | | 1.029 | 2.76 | | 0.006 | 1.344 | | 5.740 | *** |
| Chronic Kidney Disease | 5.807 | | 2.463 | 4.15 | | 0.000 | 2.528 | | 13.336 | *** |
| Hydroxychloroquine | 0.323 | | 0.110 | -3.33 | | 0.001 | 0.166 | | 0.629 | *** |
| Azithromycin | 2.442 | | 0.747 | 2.92 | | 0.004 | 1.341 | | 4.448 | *** |
| Steroids | 4.776 | | 1.460 | 5.12 | | 0.000 | 2.623 | | 8.695 | *** |
| tocilizumab | 23.080 | | 8.413 | 8.61 | | 0.000 | 11.297 | | 47.153 | *** |
| Plasma | 6.949 | | 2.824 | 4.77 | | 0.000 | 3.133 | | 15.413 | *** |
| Severity scale on admission | 9.045 | | 2.577 | 7.73 | | 0.000 | 5.175 | | 15.810 | *** |
| Constant | 0.000 | | 0.000 | -10.25 | | 0.000 | 0.000 | | 0.000 | *** |
| Mean dependent var | | 0.063 | | | SD dependent var | | | 0.242 | |  |
| Pseudo r-squared | | 0.581 | | | Number of obs | | | 1791.000 | |  |
| Chi-square | | 486.381 | | | Prob > chi2 | | | 0.000 | |  |
| Akaike crit. (AIC) | | 375.399 | | | Bayesian crit. (BIC) | | | 441.286 | |  |
| Hosmer-Lemeshow chi2(8) | | 6.41 | | | area under ROC curve | | | 0.9663 | |  |
| Hosmer-Lemeshow Prob > chi2 | | 0.6015 | | | Mean VIF | | | 1.26 | |  |
| **** p<0.01, ** p<0.05, * p<0.1* | | | | | | | | | |  |

### Secondary outcome analysis : requirement for oxygenation on presentation

**Logistic regression: Crude**

| oxygenation on admission | Coef. | | St.Err. | t-value | | p-value | [95% Conf | | Interval] | Sig |
| --- | --- | --- | --- | --- | --- | --- | --- | --- | --- | --- |
| Sickle cell disease | 0.856 | | 0.457 | -0.29 | | 0.770 | 0.301 | | 2.435 |  |
| Constant | 0.137 | | 0.010 | -27.09 | | 0.000 | 0.119 | | 0.159 | *** |
| Mean dependent var | | 0.121 | | | SD dependent var | | | 0.326 | |  |
| Pseudo r-squared | | 0.000 | | | Number of obs | | | 1792.000 | |  |
| Chi-square | | 0.089 | | | Prob > chi2 | | | 0.766 | |  |
| Akaike crit. (AIC) | | 1322.791 | | | Bayesian crit. (BIC) | | | 1333.773 | |  |
| **** p<0.01, ** p<0.05, * p<0.1* | | | | | | | | | |  |

**Logistic regression : Multivariate**

| oxygenation on admission | Coef. | | St.Err. | t-value | | p-value | [95% Conf | | Interval] | Sig |
| --- | --- | --- | --- | --- | --- | --- | --- | --- | --- | --- |
| Sickle cell disease | 1.389 | | 0.851 | 0.54 | | 0.592 | 0.418 | | 4.615 |  |
| Age | 1.036 | | 0.007 | 4.96 | | 0.000 | 1.022 | | 1.051 | *** |
| male | 1.125 | | 0.230 | 0.58 | | 0.563 | 0.754 | | 1.680 |  |
| Bahraini Nationality | 0.479 | | 0.099 | -3.57 | | 0.000 | 0.319 | | 0.718 | *** |
| Hypertension | 1.217 | | 0.270 | 0.89 | | 0.375 | 0.788 | | 1.879 |  |
| Chronic kidney disease | 2.591 | | 0.903 | 2.73 | | 0.006 | 1.309 | | 5.128 | *** |
| COPD | 7.628 | | 7.931 | 1.95 | | 0.051 | 0.994 | | 58.536 | * |
| symptoms | 1.835 | | 0.619 | 1.80 | | 0.072 | 0.947 | | 3.555 | * |
| fever | 2.073 | | 0.389 | 3.89 | | 0.000 | 1.436 | | 2.994 | *** |
| cough | 1.659 | | 0.343 | 2.45 | | 0.014 | 1.107 | | 2.487 | ** |
| sob | 6.861 | | 1.288 | 10.26 | | 0.000 | 4.749 | | 9.912 | *** |
| Heart rate | 1.025 | | 0.006 | 4.03 | | 0.000 | 1.013 | | 1.038 | *** |
| Diastolic blood pressure | 0.974 | | 0.008 | -3.32 | | 0.001 | 0.959 | | 0.989 | *** |
| Constant | 0.004 | | 0.004 | -6.15 | | 0.000 | 0.001 | | 0.023 | *** |
| Mean dependent var | | 0.121 | | | SD dependent var | | | 0.326 | |  |
| Pseudo r-squared | | 0.290 | | | Number of obs | | | 1708.000 | |  |
| Chi-square | | 364.810 | | | Prob > chi2 | | | 0.000 | |  |
| Akaike crit. (AIC) | | 920.745 | | | Bayesian crit. (BIC) | | | 996.948 | |  |
| Hosmer-Lemeshow chi2(8) | | 8.34 | | | area under ROC curve | | | 0.8662 | |  |
| Hosmer-Lemeshow Prob>chi2 | | 0.4013 | | | Mean VIF | | | 1.3 | |  |
| **** p<0.01, ** p<0.05, * p<0.1* | | | | | | | | | |  |
